# Is attachment style in early childhood associated with mental health difficulties in late adolescence?

**DOI:** 10.1101/2021.03.22.21254117

**Authors:** Philippa Clery, Angela Rowe, Marcus R. Munafò, Liam Mahedy

**Author notes:** **Corresponding Author** Dr Philippa Clery, School of Psychological Science, University of Bristol, 12a Priory Road, Bristol, BS8 1TU, UK.

## Abstract

**Background:** Although existing research has suggested an association between insecure attachment styles and mental health difficulties, these studies often have small sample sizes, use cross-sectional designs, and measure attachment as a discrete variable at a single point. It is also unclear whether these associations persist into late adolescence. In this prospective study we aimed to determine whether insecure attachment style in early childhood is associated with depression and self-harm at 18 years.

**Methods:** We used data from the Avon Longitudinal Study of Parents and Children cohort. Mothers completed attachment related questionnaires when their child was 18, 30, and 42 months old. Offspring depression and lifetime self-harm was assessed at 18 years using the Clinical Interview Schedule-Revised. Attachment was derived as a latent variable in a structural equation modelling framework. Logistic regression was performed on participants with complete attachment data (n=7032) to examine the association between attachment style and depression and self-harm, with adjustment for potential confounders. Differential dropout was accounted for using multiple imputation.

**Results:** We found some evidence for an association between attachment quality and depression and self-harm. In the fully adjusted imputed model, a one standard deviation increase in insecure attachment was associated with a 13% increase in the odds of depression at age 18 (OR=1.13; 95%CI=1.00 to 1.27) and a 14% increase in the odds of self-harm at age 18 (OR=1.14; 95%CI=1.02 to 1.25).

**Conclusions:** Our findings strengthen the evidence suggesting that an insecure attachment style assessed in early childhood is associated with mental health difficulties in late adolescence. If this association is causal, policies to support parenting or attachment-based interventions to improve attachment quality could help reduce mental health difficulties in adolescence/young adulthood.

## Introduction

Rates of depression and hospital presentations with self-harm amongst children and adolescents are rising (Griffin et al., 2018; Sadler et al., 2018). Identifying factors that contribute to mental health difficulties in young people as early as possible should be a priority to inform prevention strategies. One area of interest is the formation of attachment styles in the early caregiving environment. Attachment theory (Bowlby, 1969) offers a framework for understanding psychopathology development (Mikulincer & Shaver, 2012). Through repeated early childhood interactions with primary caregivers, children develop an ‘internal working model’ of themselves and others, which influences emotional and behavioural regulation across the lifespan (Cassidy & Shaver, 2016). Extensive evidence suggests that attachment style is a key predictor of emotion regulation, coping strategies, adjustment to stressors and therefore vulnerability to mental health problems (Bosmans et al., 2020; Mikulincer & Shaver, 2012). Children who are more securely attached (i.e., experience responsive caregiving that provides an emotional secure base) see others as responsive in times of need, form more positive peer relationships, and regulate their emotions more effectively (Thompson, 2008; Weinfeld, Sroufe, & Egeland, 2008). In contrast, those who are insecurely attached (i.e., experience insensitive or inconsistent caregiving) develop poor affect regulation, report lower coping abilities and an increased vulnerability to perceived stressors (Mikulincer & Shaver, 2008).

Decades of attachment research has tried to clarify the association between attachment style and psychopathology. However, findings are inconsistent and limited by methodological problems. Some recent meta-analyses have found small-to-moderate associations between insecure attachment styles and internalising problems (Madigan et al., 2016, Spruit et al., 2020), whilst others have only found an association between the insecure-avoidant attachment style and internalising problems (Groh, Roisman, Ijzendoorn, Bakermans-Kranenburg, & Fearon, 2012; Madigan, Atkinson, Laurin, & Benoit, 2013) or not at all (Brumariu & Kerns, 2010). Inconsistences could be due to the use of small sample sizes, short follow-up periods, cross-sectional designs, and different attachment measures.

Most studies are limited to either childhood *or* adulthood samples, thereby failing to bridge the period between childhood attachment and adolescent/adult psychopathology. Studies using *early* childhood attachment measures are deemed most appropriate for evaluating the central tenet of attachment theory (i.e., that *early* attachments influence psychological development) (Groh, Fearon, IJzendoorn, Bakermans-Kranenburg, & Roisman, 2017). The Strange Situation Procedure (SSP) (Ainsworth, Blehar, Waters, & Wall, 1978) is considered the gold-standard for measuring childhood attachment (Crittenden, Robson, & Tooby, 2015), and classifies a child’s attachment style (secure, insecure-avoidant or insecure-resistant) based on observed behaviours of the child in a situation where their caregiver returns after leaving them with a stranger for a short period. However, existing studies using the SSP typically have small sample sizes (median n=52), short follow-up durations (median=18 months), measure attachment security at one time-point only, and use behavioural outcomes reported by parents rather than child self-reports (see Madigan et al., 2013). Furthermore, categorical attachment styles can be problematic (Brumariu & Kerns, 2010) as variation in attachment patterns is thought to be largely continuous (Fraley & Spieker, 2003). Research into adolescent and adult attachment addresses more clinically relevant mental health outcomes; however, these studies are also limited by small sample sizes, the use of cross-sectional designs (see Spruit et al., 2020), or use proxy adult romantic relationship attachment measures (e.g., Eberhart & Hammen, 2006).

In an effort to strengthen current evidence, we used data from the UK Avon Longitudinal Study of Parents and Children (ALSPAC) birth cohort study to examine whether attachment quality, assessed repeatedly at ages 18, 30, and 42 months via maternal report, is prospectively associated with offspring reports of mental health assessed at 18 years. We used a structural equation modelling (SEM) approach to derive a continuous attachment measure, in order to increase statistical power and reduce measurement error. We also adjusted for relevant confounders, used a long follow-up period, and included both maternal and offspring reports. Our overall aim was to estimate the magnitude of any prospective association between a more insecure attachment style and (1) a diagnosis of depression defined by International Classification of Diseases (ICD)-10 criteria (World Health Organisation, 1992) and/or (2) self-harm, at 18 years.

## Methods

### Study design and participants

The sample comprised participants from ALSPAC, a transgenerational ongoing prospective UK cohort study which examines the influences on health and development across the lifespan. Pregnant women resident in the former Avon Health Authority in South-West England with expected delivery dates 1/4/1991 to 31/12/1992 were invited to take part. The initial number of pregnancies enrolled is 14,541 (for these, at least one questionnaire was returned or a ‘Children in Focus’ clinic was attended by 19/7/1999). Of these initial pregnancies, there was a total of 14,676 fetuses, resulting in 14,062 live births and 13,988 children who were alive at one year. When the oldest children were seven years old, an attempt was made to bolster the sample with eligible cases who had failed to join the study originally. As a result, when considering variables collected from age seven onwards there are data available for more than 14,541 pregnancies. The number of new pregnancies not in the initial samples (known as phase I enrolment) that are currently represented on the built files and reflecting enrolment status at the age of 24, is 913 (456, 262 and 195 recruited during phases II, III and IV respectively). The total sample size for analyses using any data collected after age seven is therefore 15,454 pregnancies, resulting in 15,589 fetuses. Of these, 14,901 were alive at one year. The phases of enrolment are described in more detail in the cohort profile paper and its update (Boyd et al., 2013; Fraser et al., 2013; Northstone et al., 2019).

A 10% sample of the ALSPAC cohort, known as the Children in Focus (CiF) group, attended clinics at the University of Bristol at various time intervals between four and 61 months of age. The CiF group were chosen at random from the last six months of ALSPAC births (1432 families attended at least one clinic). Excluded were those mothers who had moved out of the area or were lost to follow-up, and those partaking in another study of infant development in Avon. Parent and child participants have been regularly followed-up since recruitment through postal questionnaires and attending research clinics. Data collected from participants at 22 years and onwards used REDCap (Research Electronic Data Capture) electronic data capture tools hosted at the University of Bristol. REDCap is a secure, web-based software platform (Harris et al., 2009). The ALSPAC study website contains details of all data, available through a fully searchable data dictionary and variable search tool (http://www.bristol.ac.uk/alspac/researchers/our-data/).

This current study used a sample of 14,732 participants in the core ALSPAC dataset. For reasons of confidentiality, data for children of triplet and quadruplet pregnancies were removed. We used data from 11,741 participants who returned questionnaires for exposure variables. Analysis was based on 4287 adolescents who completed questionnaires on depression and self-harm at the ‘Teen-Focus-4’ clinic at 18 years.

Ethics approval was obtained from the ALSPAC Law and Ethics committee and the Local Research Ethics Committees. Informed consent for the use of data collected via questionnaires and clinics was obtained from participants following the recommendations of the ALSPAC Ethics and Law Committee at the time.

### Measures

#### Exposure: Attachment style

Mothers completed questionnaires when their child was 18, 30 and 42 months old, including a question based on the SSP: “When you and your toddler meet again after being apart for an hour or more, how often (always/usually, sometimes or hardly ever) does she/he (a) move away or avoid looking at you, (b) push you away (c) run to you for a hug or cuddle?”. A fourth response “we are never apart” was included in the 18 and 42 month questionnaires, but was not available for all three time-points and does not correspond to an attachment style, so was not included in analyses. Descriptive information on attachment measures is in Table S1. A latent variable model was created to combine the three attachment questions across the three time-points, to model the construct of attachment style as a continuous variable. Higher values indicate more insecure attachment. Mothers who completed all attachment questions across all time-points (n=7032) were included in regression analysis.

#### Outcomes: Depression and Self-harm

At the 18-year clinic (M=17.8, SD=0.46), participants completed a self-administered computerised Clinical Interview Schedule-Revised (CIS-R) (Lewis, Pelosi, Araya, & Dunn, 1992). The CIS-R diagnoses depression according to the ICD-10 criteria (World Health Organisation, 1992). A binary variable of depression (depressed or not) was used to record those who met these criteria (n=338). The CIS-R is designed for, and has been widely used in, community samples as well as ALSPAC studies (e.g. Bowes et al., 2015), and is a reliable and valid measurement of depression (Bebbington et al., 2003). There is close agreement between the self-administered computerised and interviewer administrated versions (Patton et al., 1999).

The CIS-R also provides a binary ‘self-harm indicator’ variable derived from the question “Have you ever hurt yourself on purpose in any way (e.g., by taking an overdose of pills or by cutting yourself)?” Response options were “yes” or “no”. Participants were classified as having a life-time history of self-harm if they answered “yes”. The wording of this question is taken from the Childhood Interview for DSM-IV Borderline Personality Disorder (Zanarini, 2003). All self-harm measures in ALSPAC use this screening question, which is widely used in research (e.g., Kidger, Heron, Lewis, Evans, & Gunnell, 2012).

#### Confounders

We identified a range of possible confounders of the relationship between attachment style and depression and self-harm (Russell et al., 2019; Spruit et al., 2020). These were taken from maternal self-report questionnaires early in the life of the cohort, either during pregnancy or at eight months post-natal, to ensure confounders were measured before exposure and to reduce missingness. Confounders included: sex; socioeconomic positioning assessed using housing tenure (home ownership status in three categories of owned/mortgaged, private rented, council/housing association rented) and parental social class (indicating the highest paternal or maternal occupation from higher managerial/professional to unskilled work); maternal education (mother’s highest educational qualification from high to low: ‘degree’ (university), ‘A-levels’ (school assessments aged 18), ‘O-level’ (school assessments aged 16), ‘vocational’ (occupational skills), ‘Certificate of Secondary Education’, ‘none’); maternal depression (Edinburgh Post-natal Depression Score at eight months – linear scale from 0 to 30, where higher values indicate worse depression (Cox, Holden, & Sagovsky, 1987)); maternal alcohol usage (in five categories of ‘never’, ‘<1 glass-per-week’, ‘>1 glass-per-day’, ‘1-2 glasses-per-day’ and ‘≥3 glasses-per-day’); maternal smoking (yes/no); number of other children in the household under the age of 15; and parental relationship, including a score of partner affection (a continuous scale from 0 to 49, where higher scores indicate higher levels of affection) and whether parents were separated or divorced (yes/no). Figure 1 shows the timeline of data collection for exposure, outcome and confounding variables.

**Fig 1:**
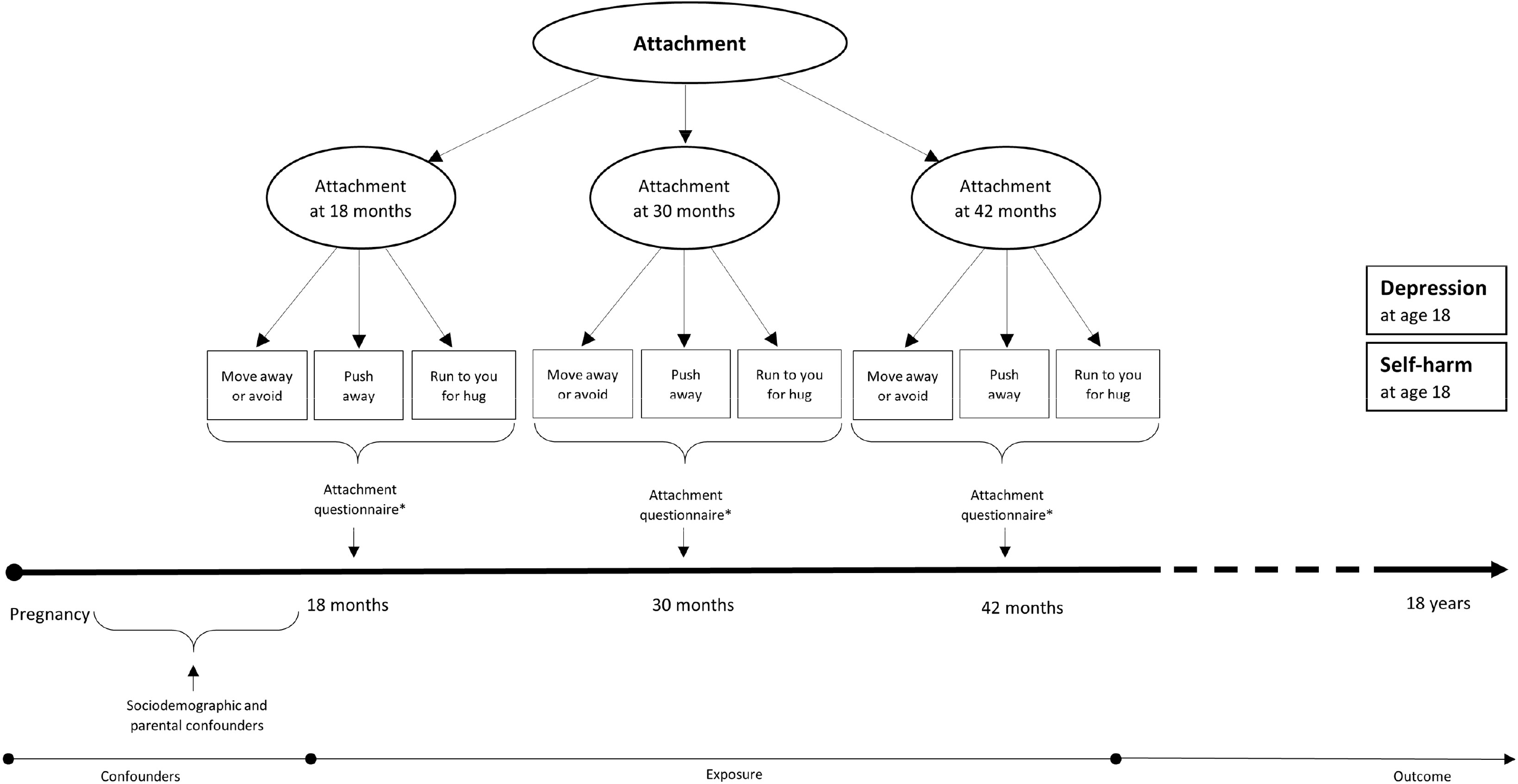
Timeline for data collection of possible confounders, exposure and outcome data.

### Statistical analysis

#### Childhood attachment

SEM was used with the ‘lavaan’ package in R (Rosseel, 2012) to create a second order latent variable model to model a continuous construct of attachment across three time-points in early childhood (using nine observed variables in total; three variables each measured at 18, 30 and 42 months). SEM was utilised using the diagonally weighted least squares (DWLS) to estimate the model parameters, and the full weight matrix to compute robust standard errors and a mean- and variance-adjusted test statistic (Rosseel, 2012). Missing data in the model were dealt with using pairwise deletion, which calculates polychoric correlations for each pair of variables – using all available cases that provided data on both variables – which are used in the DWLS fit function (Katsikatsou, Moustaki, Yang-Wallentin, & Jöreskog, 2012). The fit of the SEM model was assessed using the root-mean-square error of approximations (RMSEA) (Steiger, 1990), Comparative Fit Index (CFI) (Bentler, 1990), and Tucker Lewis Index (TLI) (Tucker & Lewis, 1973). Goodness of fit was indicated by an RMSEA of less than 0.06 (Hu & Bentler, 1999). As a next step, logistic regression modelling using probit models was used to investigate the association between attachment and depression/self-harm at 18 years. Models were adjusted for different levels of confounders. Results are presented as odds ratios and their 95% confidence intervals and are interpreted as: each standard deviation increase in the exposure is associated with the corresponding increase/decrease in odds of having the outcome compared to not. Statistical analysis was conducted in R, version 4.0.3 (R Core Team, 2020).

#### Missing data

Using complete case data without taking missing data into account can result in biased estimates (Spratt et al., 2010). Assuming Missing at Random (MAR), we examined possible effects of missing data using an approach to impute missing data using multivariate imputation by chained equations using the ‘mice’ package in R (Buuren & Groothuis-Oudshoorn, 2011). A number of auxiliary variables predictive of incomplete variables were included: sociodemographic variables; IQ; and reports of depression and self-harm at additional ages (15-24 years) from the CIS-R in clinics and the short Moods and Feelings questionnaire (sMFQ; Angold et al., 1995). A final set of adjusted models was created for each individual, mother and parental relationship factors, on complete cases (n=2397). Multiple imputation was based on 7032 participants who had information on all of the attachment variables. Of these, n=2956 had outcome data. To ensure accurate representation of the data, 50 imputed datasets were derived. Imputation models contained all exposure, confounding and outcome data. We present a comparison of estimates from the complete case and imputed data analysis in the results.

## Results

### Descriptive statistics

Of the core ALSPAC sample (n=14,732), 11,741 (79.7%) participants had data on at least one exposure variable (i.e., answered at least one of nine attachment questions; Table S1). SEM was conducted on this sample to derive the latent classes. Figure 2 (and Table S2) show the factor loadings of attachment latent variables, which range from 0.87 to 1.00 for the first order model, and 0.97 to 1.29 for the second order model. The second order model provided an adequate fit/description of the data (RMSEA=0.04, 90%CI=0.04 to 0.04; CFI=0.96; TLI=0.95).

**Fig 2:**
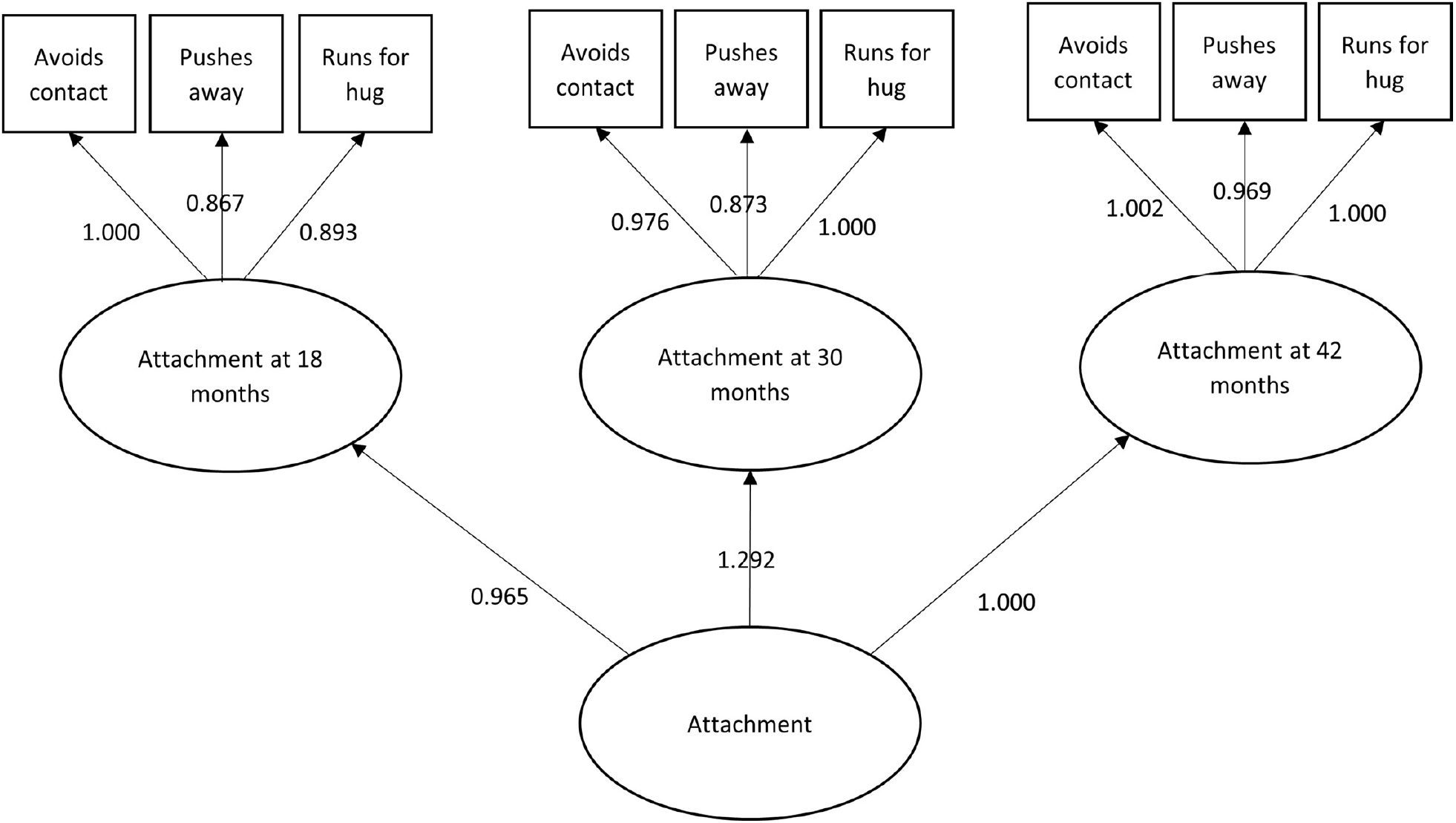
Structural equation model depicting latent variables with factor loadings

Of the core ALSPAC sample, n=9557 (64.87%) participants were invited to clinic and n=4878 (33.1%) attended. Figure S1 shows the participant flowchart, while Table S3 shows the differences between those who had data available from clinic compared to those who did not. Clinic non-attendance was associated with lower socioeconomic positioning based on housing tenure (OR=2.91, 95%CI=2.59 to 3.26) and parental occupation (OR=3.48, 95%CI=2.18 to 5.56), lower levels of maternal education (OR=4.21, 95%CI=3.67 to 4.82), parental separation or divorce (OR=1.55, 95%CI=1.32 to 1,83), and more children in the household (OR=1.18, 95%CI=1.13 to 1.23). Of the clinic participants with CIS-R data (n=4287), 338 (7.88%) had an ICD-10 diagnosis of depression and 687 (16.0%) reported life-time self-harm.

### Association between childhood attachment and adolescent depression

There was weak evidence for an association between attachment in early childhood and an ICD-10 diagnosis of depression at age 18 (Table 1). In the fully adjusted imputed model, every one standard deviation increase in insecure attachment in early childhood was associated with a 13% increase in the odds of diagnosis of depression at age 18 (OR=1.13; 95%CI=1.00 to 1.27).

**Table 1:** Associations between attachment security and depression and self-harm at age 18 (n=7032).

|  | Depression |  | Self-Harm |  |
| --- | --- | --- | --- | --- |
|  | OR (95% CI) | p-value | OR (95% CI) | p-value |
| **Model 1** | 1.09 (0.98, 1.20) | 0.12 | 1.11 (1.02, 1.20) | 0.03 |
| **Model 2** | 1.13 (1.01, 1.24) | 0.04 | 1.16 (1.06, 1.25) | <0.01 |
| **Model 3** | 1.14 (1.03, 0.12) | 0.03 | 1.17 (1.07, 1.27) | <0.01 |
| **Model 4** | 1.15 (1.01, 1.28) | 0.05 | 1.14 (1.03, 1.25) | 0.02 |
| **Model 5** | 1.13 (1.00, 1.27) | 0.08 | 1.14 (1.02, 1.25) | 0.03 |

Note: **Model 1** = unadjusted; **Model 2** = adjusted for sex; **Model 3** = further adjusted for parental social class, housing tenure, maternal education; **Model 4** = further adjusted for maternal age, maternal depression, maternal smoking and alcohol consumption in pregnancy, number of children <15 years-of-age in household; **Model 5** = further adjusted for parental affection score and whether parents separated/divorced.

There was a similar pattern of results in complete case analyses (Table S4). For every one standard deviation increase in insecure attachment, there was a 14% increase in odds in depression in the fully adjusted model (OR=1.14; 95%CI=0.94 to 1.34). However, the reduced sample size resulted in wider confidence intervals.

### Association between childhood attachment and adolescent self-harm

There was weak evidence for an association between attachment in early childhood and lifetime self-harm at age 18 (Table 1). In the fully adjusted imputed model, every one standard deviation increase in insecure attachment in early childhood was associated with a 14% increase in the odds of self-harm at age 18 (OR=1.14; 95%CI=1.02 to 1.25).

Complete case analysis results (Table S4) found that, for every one standard deviation increase in insecure attachment, there was a 5% increase in odds in self-harm in the fully adjusted model (OR=1.05; 95%CI=0.88 to 1.22)

## Discussion

### Main findings

We found some evidence for an association between a more insecure attachment style in childhood, and depression and self-harm at 18 years. In fully adjusted analysis, the odds of lifetime self-harm and diagnosis of depression at 18 years were 14% and 13% higher respectively, for children who had more insecure attachment in early childhood, compared with children who had more secure attachment.

### Comparison with literature

To our knowledge, this is the largest longitudinal study assessing the prospective relationship between attachment in pre-school-age childhood and mental health difficulties in late adolescence. Our findings are consistent with meta-analyses that show weak-to-moderate effect sizes for the association between insecure attachment and internalising symptoms (Groh et al., 2012; Madigan et al., 2013, 2016; Spruit et al., 2020). However, individual studies in meta-analyses have small sample sizes. This study’s sample size (n=7032) is larger than total combined sample sizes of the meta-analyses assessing attachment in early childhood; for example, Groh et al (2012) had a sample of n=4614, and Madigan et al (2013) n=5236.

Our effect estimates are smaller and weaker compared to some studies with larger samples (e.g. McCann, Higgins, Perra, McCartan, & McLaughlin, 2014), but these used cross-sectional designs and non-childhood attachment measures. Without access to rich data on parental and environmental variables, these studies also lack potentially relevant confounders, which our study was able to adjust for. Our results are similar to those from the largest (n=1313) longitudinal study to date (Branje, Hale, Frijns, & Meeus, 2010), where parental relationship quality only weakly predicted later depressive symptoms. However, their follow-up duration was four years, compared to ours of 14.5 years. Commonly, smaller effect sizes are reported in studies with longer duration between attachment quality and outcome measures, and where the child is younger when attachment style was measured (Spruit et al., 2020). Compared to other studies that measure attachment in early childhood, ours has the advantage of repeated attachment measures and a longer follow-up period; the current eldest age at follow-up is 11 years (Bar-Haim, Dan, Eshel, & Sagi-Schwartz, 2007), whereas ours is 18 years.

A possible explanation for the smaller effect size we observed is that early attachment may be less relevant for developing psychopathology later in adolescence. There is evidence that attachment styles are malleable into adulthood (Fraley, 2002) and that a change in attachment trajectory correlates with a change in depressive trajectory (Khan, Fraley, Young, & Hankin, 2019). Therefore, changes in relational environment and life events can alter the attachment model and have direct effects on mental health.

Looking specifically at self-harm outcomes, our findings agree with results from small, cross-sectional studies with university student samples, showing that those who engage in self-harm report a poorer quality of attachment to parents compared to those who do not (Buckmaster, McNulty, & Guerin, 2019). The shared feature of emotional dysregulation found with both insecure attachment and self-harm (Gratz & Roemer, 2008) offers a possible explanation: when faced with stress, individuals with emotion dysregulation may be less able to tolerate difficult emotions, and more likely to internalise distress, subsequently releasing it in the form of self-injuring behaviour (Ross & Heath, 2002). Another explanation is that internalising symptoms mediate this effect, lying on the causal pathway between emotional dysregulation and self-harm (Kranzler, Fehling, Anestis, & Selby, 2016). Either way, if these associations are causal, attachment style appears to be a key factor.

Given that adolescent depression and self-harm are priority areas for policymakers, efforts should focus on preventing or improving insecure attachment styles. If the associations we observe are causal, parenting interventions could help children develop secure attachment styles. Attachment quality can also be improved through attachment-based interventions by strengthening mental models of attachment security so they are more readily accessible and activated (Carnelley & Rowe, 2007). Attachment-based family therapy has been used to good effect for depression in psychiatric inpatient adolescent samples (Devacht, Bosmans, Dewulf, Levy, & Diamond, 2019).

### Strengths and Limitations

This is the largest longitudinal study to examine the prospective association between attachment in pre-school-age childhood and depression and self-harm in late adolescence. We used a structural equation modelling approach to derive a childhood attachment as a continuous latent variable measured across three occasions, providing a more robust measure than a snapshot of categorical attachment styles at one time-point. We also used outcome measures with differential respondents (maternal reports of attachment and offspring reports of depression/self-harm) that are clinically relevant. Due to its longitudinal and rich data collection, ALSPAC has a wealth of data available to adjust for confounders and perform imputation to address potential bias due to attrition.

However, our findings should be interpreted in the context of some limitations. First, the ALSPAC cohort suffers from attrition, which is higher for those who are socially disadvantaged (Wolke et al., 2009). Although it is not possible to test the MAR assumption, it was made more plausible as socioeconomic variables were found to predict clinic attendance. Adolescents who attended the 18-year clinic were more likely to come from higher socioeconomic backgrounds (Table S3). To address potential response-attrition bias, multiple imputation was performed. Findings were consistent across complete case and multiple imputation analyses. Second, we cannot be certain that the associations we observed reflect causal pathways. We also did not adjust for maternal genetics, so cannot exclude the role of shared genetic vulnerability for offspring. The longitudinal nature of the study suggests we can exclude reverse causality. However, the relationship between attachment and psychopathology could be explained, at least in part, by gene-environment correlation (Sallis et al., 2020). Future studies could address this by using Genome Wide Association Study data to conduct Mendelian randomisation analyses, which – if trio generational data were available – could also include an inter-generational component as there is evidence that the influence of parenting behaviours on offspring’s mental health transcends generations (Mahedy et al., 2014). Third, whilst sophisticated SEM methods were used to create a robust, continuous measure of attachment, the model is not based on a validated scale. The questions are similar to observations used in the SSP (Ainsworth et al., 1978), but in the SSP the child is left with a stranger, whereas we do not know who the child was left with in this study. We also did not examine the categories that SSP is designed to elicit. However, it is now considered that measuring attachment on continuous dimensions is more representative (Fraley & Spieker, 2003). Further, questions were focussed on mothers as primary caregivers, without considering fathers. However, there is evidence that psychopathology is more strongly associated with maternal bonding than with paternal bonding (Enns, Cox, & Clara, 2002). Traditionally, mothers are the primary emotional caregiver and their parenting behaviour may be more influential (Cinamon & Rich, 2002).

## Conclusion

Using rich longitudinal data from a large UK birth cohort our findings add to the existing literature by providing evidence that an insecure attachment style developed in early childhood is associated with depression and self-harm in late adolescence. If the results we observe are causal, there is potential importance of the *early* caregiving environment for influencing attachment style development, and in turn, mental health. To target adolescent mental health difficulties, policies and interventions could support parenting behaviours that foster the development of secure attachment styles, or attachment-based therapies could be used to improve attachment quality.

## Data Availability

Data are available upon request to author.

**Key points and relevance**

### What’s known?

- Attachment style influences a child’s interpersonal relationships, emotion regulation, and development of mental health difficulties.
- Previous studies looking at the association between attachment and psychopathology are limited by small sample sizes, cross-sectional designs, and poor attachment measures.

### What’s new?

- This large, prospective study in ALSPAC, using a continuous measure of repeated attachment measures in early childhood and clinically relevant outcomes at age 18, suggests an association between more insecure attachment and depression and self-harm.

### What’s relevant?

- To target adolescent mental difficulties, policies and interventions could support parenting behaviours to create an early caregiving environment that fosters the development of a secure attachment style, or attachment-based therapies could improve attachment quality.

## Acknowledgements

We are extremely grateful to all the families who took part in this study, the midwives for their help in recruiting them, and the whole ALSPAC team, which includes interviewers, computer and laboratory technicians, clerical workers, research scientists, volunteers, managers, receptionists and nurses.

## Authorship

LM conceptualised the study. LM and PC designed the study and performed data analysis and interpretation. PC wrote the manuscript. All authors were involved in manuscript drafts and revisions, and approve the final version.

## Abbreviations

ALSPAC: Avon Longitudinal Study of Parents and Children
SEM: Structural Equation Modelling

