## Supplementary Figures and Tables for "Is attachment style in early childhood associated with mental health difficulties in late adolescence?"

**Table S1:** Attachment variables, corresponding to the three questions asked at the three different ages. This includes the participants who answered with “never apart”. Sample is based on the core ALSPAC sample (n=14,732).

| | Question 1: Avoids contact with mother/carer n (%) | | | | Question 2: Pushes mother/carer away n (%) | | | | Question 3: Runs for hug n (%) | | | |
|---|---|---|---|---|---|---|---|---|---|---|---|---|
| **Age (months)** | Hardly ever | Sometimes | Always/ usually* | Never apart | Hardly ever | Sometimes | Always/ usually* | Never apart | Always/ Usually* | Sometimes | Hardly ever | Never apart |
| **18** | 8115 (86.56) | 601 (6.41) | 61 (0.65) | 598 (6.38) | 8233 (88.1) | 474 (5.07) | 43 (0.46) | 598 (6.40) | 8947 (81.2) | 1283 (11.7) | 188 (1.71) | 598 (5.43) |
| **30** | 9708 (94.9) | 505 (4.94) | 15 (0.15) | n/a** | 9681 (94.7) | 529 (5.18) | 9 (0.09) | n/a** | 7695 (75.5) | 2380 (23.3) | 124 (1.22) | n/a** |
| **42** | 9564 (95.68) | 328 (3.28) | 5 (0.05) | 99 (0.99) | 9571 (95.75) | 318 (3.18) | 8 (0.08) | 99 (0.99) | 7505 (75.1) | 2257 (22.6) | 135 (1.35) | 99 (0.99) |

Note: * on 18 months questionnaire, the category ‘usually’ was asked, on the 30 and 42 months questionnaire it was re-worded to ‘always’; ** “never apart” was not an option on the questionnaire at 30 months; categories were collapsed into the above one if the number of observations was <15.

**Table S2:** Structural equation modelling of attachment (standardised coefficients)

| **Latent Variables** | | |
|---|---|---|
| **First order model** | **Questionnaire item*** | **DWLS** |
| Attachment at 18 months | | |
| | Frequency infant avoids contact with mother after separation | 1.000 |
| | Frequency infant pushes mother away after separation | 0.867 |
| | Frequency infant cuddles mother after separation** | 0.893 |
| Attachment at 30 months | | |
| | Frequency infant avoids contact with mother after separation | 0.976 |
| | Frequency infant pushes mother away after separation | 0.873 |
| | Frequency infant cuddles mother after separation** | 1.000 |
| Attachment at 42 months | | |
| | Frequency infant avoids contact with mother after separation | 1.002 |
| | Frequency infant pushes mother away after separation | 0.969 |
| | Frequency infant cuddles mother after separation** | 1.000 |
| **Second order model** | **First order model latent variable** | |

| | | |
|---|---|---|
| Attachment over the three ages | | |
| | Attachment at 18 months | 0.965 |
| | Attachment at 30 months | 1.292 |
| | Attachment at 42 months | 1.000 |

**Note**: * Based on questions: when you and your toddler meet again after being apart for an hour or more, how often ((a) always/usually, (b) sometimes, or (c) hardly ever) do they: (1) move away or avoid looking at you (2) push you away (3) run to you for a hug or cuddle?; **reverse coded so remains consistent with other measures that higher scores = poor attachment; DWLS estimate = diagonally weighted least squares.

**Fig S1**: Flowchart of ALSPAC participants showing attrition.

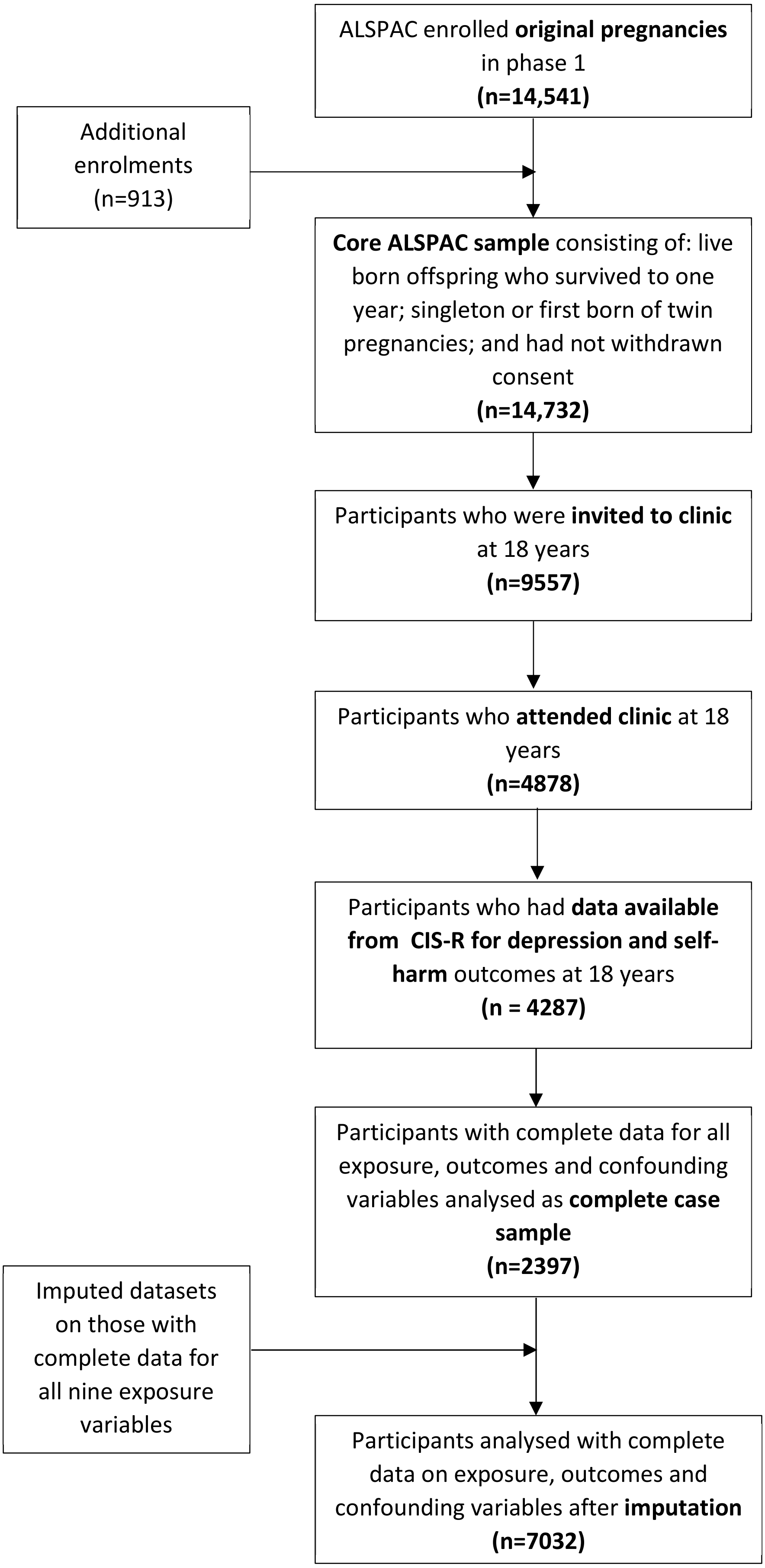

**Table S3** : Selective attrition for participants who attended the 18-year clinic.

| | | Available/ attended clinic (n= 4878*) | Not available/ did not attend clinic (n= 9854*) | OR (95% CI) |
|---|---|---|---|---|
| | | n (%) | n (%) | |
| **Gender** | | | | |
| | Female | 2743 (56.2) | 4079 (41.4) | 0.61 (0.57, 0.66) |
| **Housing tenure** | | | | |
| **High** | Owned/mortgaged | 3978 (81.5) | 5894 (59.8) | Ref |
| | Private rented | 216 (4.43) | 762 (7.73) | 2.38 (2.04, 2.78) |
| **Low** | Council/housing association rented | 408 (8.36) | 1757 (17.8) | 2.91 (2.59, 3.26) |
| **Parental social class** | | | | |
| **High** | I | 800 (16.4) | 738 (7.49) | Ref |
| | II | 2055 (42.1) | 2773 (28.1) | 1.46 (1.30, 1.64) |
| | III (non-manual) | 1042 (21.4) | 1904 (19.3) | 1.98 (1.75, 2.25) |
| | III (manual) | 408 (8.36) | 1155 (11.7) | 3.07 (2.64, 3.57) |
| | IV | 162 (3.32) | 421 (4.27) | 2.82 (2.29, 3.46) |
| **Low** | V | 24 (0.49) | 77 (0.78) | 3.48 (2.18, 5.56) |
| **Maternal education** | | | | |
| **High** | Degree | 872 (17.9) | 737 (7.48) | Ref |
| | A level | 1308 (26.8) | 1485 (0.15) | 1.34 (1.19, 1.52) |
| | O level | 1611 (33.0) | 2712 (27.5) | 1.99 (1.77, 2.24) |
| | Vocational | 355 (7.28) | 874 (8.87) | 2.91 (2.49, 3.41) |

| | | | | |
|---|---|---|---|---|
| **Low** | Secondary education | 554 (11.4) | 1971 (20.0) | 4.21 (3.67, 4.82) |
| **Maternal alcohol usage in pregnancy** | | | | |
| | Never | 2063 (42.3) | 3932 (39.9) | Ref |
| | < 1 glass per week | 1958 (40.1) | 3141 (31.9) | 0.84 (0.78, 0.91) |
| | 1 glass per day | 647 (13.3) | 1186 (12.0) | 0.96 (0.86, 1.07) |
| | 1-2 glasses per day | 72 (1.48) | 140 (1.42) | 1.02 (0.76, 1.36) |
| | ≥3 glasses per day | 15 (0.31) | 27 (0.27) | 0.94 (0.50, 1.78) |
| **Maternal smoking in pregnancy** | | | | |
| | Yes | 793 (16.3) | 2570 (26.1) | 2.15 (1.97, 2.35) |
| **Parental separation or divorce** | | | | |
| | Yes | 228 (4.67) | 493 (5.00) | 1.55 (1.32, 1.83) |
| | | **Mean (SD)** | **Mean (SD)** | |
| **Maternal age** | (linear scale) | 29.7 (4.58) | 28.0 (4.9) | 0.93 (0.92, 0.94) |
| **Number of children in household < 15 years old** | (linear scale) | 1.75 (0.99) | 1.90 (0.96) | 1.18 (1.13, 1.23) |
| **Maternal depression** | (linear scale 0-29) | 5.10 (4.49) | 5.63 (4.82) | 1.02 (1.02, 1.03) |
| **Partner affection** | (linear scale 0-49) | 21.45 (7.06) | 21.72 (7.38) | 1.01 (0.99, 1.01) |

Note: *from core ALSPAC sample n=14732; highest of either parent's social class based on 5 employment categories: I = professional; II = managerial or technical, III = skilled manual or non-manual, IV = unskilled manual, V = casual and lowest grade work; maternal depression as measured by the validated scale Edinburgh Postnatal Depression Score at 8 months post-natal;

partner affection score (created from individual questions assessing affection toward partner) where higher scores indicate higher levels of affection.

**Table S4:** Associations between attachment and (a) depression (b) self-harm at age 18. Results are based on complete case analysis (n=2397).

| | **Depression** | | **Self-Harm** | |
|---|---|---|---|---|
| | OR (95% CI) | p-value | OR (95% CI) | p-value |
| **Model 1** | 1.07 (0.88, 1.26) | 0.49 | 1.02 (0.86,1.18) | 0.82 |
| **Model 2** | 1.11 (0.92, 1.31) | 0.26 | 1.08 (0.91, 1.25) | 0.37 |
| **Model 3** | 1.13 (0.93, 1.32) | 0.24 | 1.08 (0.91, 1.24) | 0.41 |
| **Model 4** | 1.13 (0.94, 1.33) | 0.21 | 1.05 (0.88, 1.23) | 0.54 |
| **Model 5** | 1.14 (0.94, 1.34) | 0.21 | 1.05 (0.88, 1.22) | 0.58 |

Note: **Model 1** = unadjusted; **Model 2** = adjusted for sex; **Model 3** = further adjusted for parental social class, housing tenure, maternal education; **Model 4** = further adjusted for maternal age, maternal depression, maternal smoking and alcohol consumption in pregnancy, number of children <15 years-of-age in household; **Model 5** = further adjusted for parental affection score and whether parents separated/divorced.
